# Tracking the Preclinical Progression of Transthyretin Amyloid Cardiomyopathy Using Artificial Intelligence-Enabled Electrocardiography and Echocardiography

**DOI:** 10.1101/2024.08.25.24312556

**Authors:** Evangelos K. Oikonomou, Veer Sangha, Sumukh Vasisht Shankar, Andreas Coppi, Harlan M. Krumholz, Khurram Nasir, Edward J. Miller, Cesia Gallegos-Kattan, Mouaz H. Al-Mallah, Sadeer Al-Kindi, Rohan Khera

## Abstract

**Background and Aims:** The diagnosis of transthyretin amyloid cardiomyopathy (ATTR-CM) requires advanced imaging, precluding large-scale pre-clinical testing. Artificial intelligence (AI)-enabled transthoracic echocardiography (TTE) and electrocardiography (ECG) may provide a scalable strategy for pre-clinical monitoring.

**Methods:** This was a retrospective analysis of individuals referred for nuclear cardiac amyloid testing at Yale-New Haven Health System (YNHHS, internal cohort) and Houston Methodist Hospitals (HMH, external cohort). Deep learning models trained to discriminate ATTR-CM from age/sex-matched controls on TTE videos (AI-Echo) and ECG images (AI-ECG) were deployed to generate study-level ATTR-CM probabilities (0-100%). Longitudinal trends in AI-derived probabilities were examined using age/sex-adjusted linear mixed models, and their discrimination of future disease was evaluated across preclinical stages.

**Results:** Among 984 participants at YNHHS (median age 74 years, 44.3% female) and 806 at HMH (69 years, 34.5% female), 112 (11.4%) and 174 (21.6%) tested positive for ATTR-CM, respectively. Across cohorts and modalities, AI-derived ATTR-CM probabilities from 7,352 TTEs and 32,205 ECGs diverged as early as 3 years before diagnosis in cases versus controls (*p*_time(x)group interaction_*≤*0.004). Among those with both AI-Echo and AI-ECG available one-to-three years *before* nuclear testing (n=433 [YNHHS] and 174 [HMH]), a double-negative screen at a 0.05 threshold (164 [37.9%] and 66 [37.9%], vs all else) had 90.9% and 85.7% sensitivity (specificity of 40.3% and 41.2%), whereas a double-positive screen (78 [18.0%] and 26 [14.9%], vs all else) had 85.5% and 88.9% specificity (sensitivity of 60.6% and 42.9%).

**Conclusions:** AI-enabled echocardiography and electrocardiography may enable scalable risk stratification of ATTR-CM during its pre-clinical course.

Structured Graphical Abstract.
Artificial intelligence (AI)-enhanced interpretation of standard echocardiographic videos and electrocardiographic (ECG) images may serve as digital biomarkers of disease progression during the early pre-clinical and clinical stages of transthyretin amyloid cardiomyopathy. We show that across two geographically distinct cohorts of individuals referred for nuclear cardiac amyloid testing, cases exhibit significantly faster progression in their AI-defined probabilities in the years before nuclear cardiac amyloid testing, compared with controls, a finding that was consistent across cohorts and modalities. These findings suggest that AI-enabled echocardiography and ECG may be able to identify those at risk for ATTR-CM up to 3 years before clinical diagnosis through standard clinical pathways. AI: artificial intelligence; ATTR-CM: transthyretin amyloid cardiomyopathy; ECG: electrocardiography; TTE: transthoracic echocardiography.

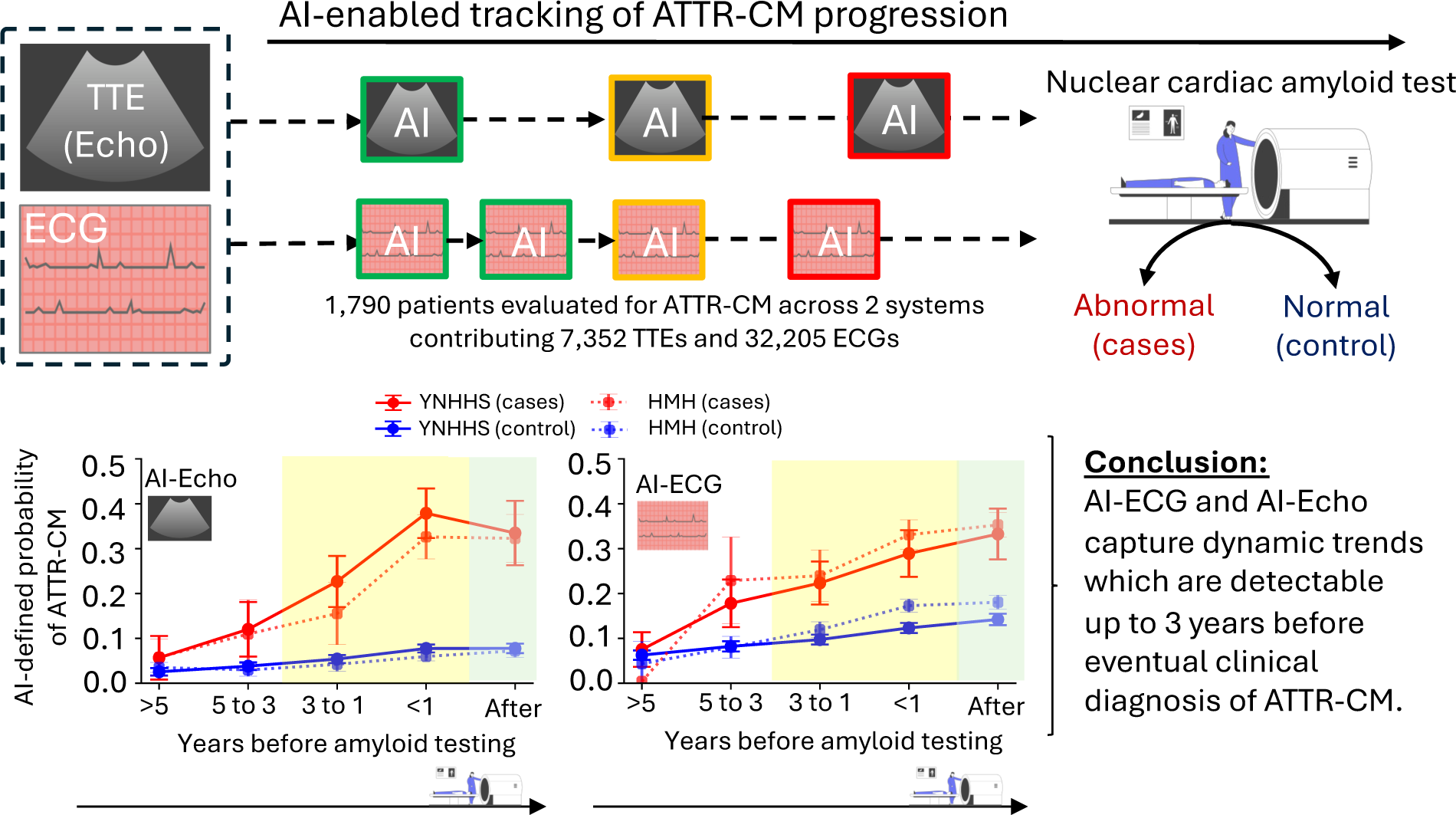

## INTRODUCTION

Awareness of the insidious onset and progression of transthyretin amyloid cardiomyopathy (ATTR-CM) is increasing, with growing recognition of its under-appreciated prevalence and links to incident heart failure and premature mortality.^1–6^ This is partly due to the evolution of several new therapeutic agents that can effectively reduce the associated morbidity and mortality.^7–9^ These therapies can stabilize abnormally folded transthyretin protein that deposits in myocardium,^7,8^ silence its production,^9–11^ and even promote its clearance,^12^ thereby modifying the course of the disease, especially when deployed early during its course.^13^ Despite these advances, on-treatment mortality and morbidity remain high,^7,8,13^ suggesting the need for earlier identification and treatment. The key challenge with optimizing the use of these new therapeutic agents is identifying individuals before the onset of symptoms, long before traditional diagnostic testing is usually performed. While dedicated nuclear cardiac amyloid testing remains a key part of the diagnostic cascade,^1,14^ the need for access to specialized centers, its cost, and radiation exposure, limit its broader use in identifying those with pre-clinical disease. Therefore, there are currently no scalable, automated strategies to identify individuals with pre-clinical disease and track its progression to flag the appropriate timing for intervention.

In this study, we hypothesized that structural, electrical, and mechanical changes induced by the deposition of amyloid fibrils are detectable through AI-enhanced interpretation of routine transthoracic echocardiography (TTE) and 12-lead electrocardiography (ECG) long before the development of clinical disease and its recognition, thus offering a scalable strategy for the early identification of pre-clinical ATTR-CM. Using a multi-center cohort of individuals referred for ATTR-CM evaluation, both with and without a confirmed diagnosis, we aimed to characterize longitudinal trajectories of quantitative AI-defined echocardiographic (AI-Echo) and electrocardiographic (AI-ECG) phenotypes and their performance as stand-alone or combined tests in distinguishing cases from controls during the years preceding diagnosis (**Figure 1**).

**Figure 1.**
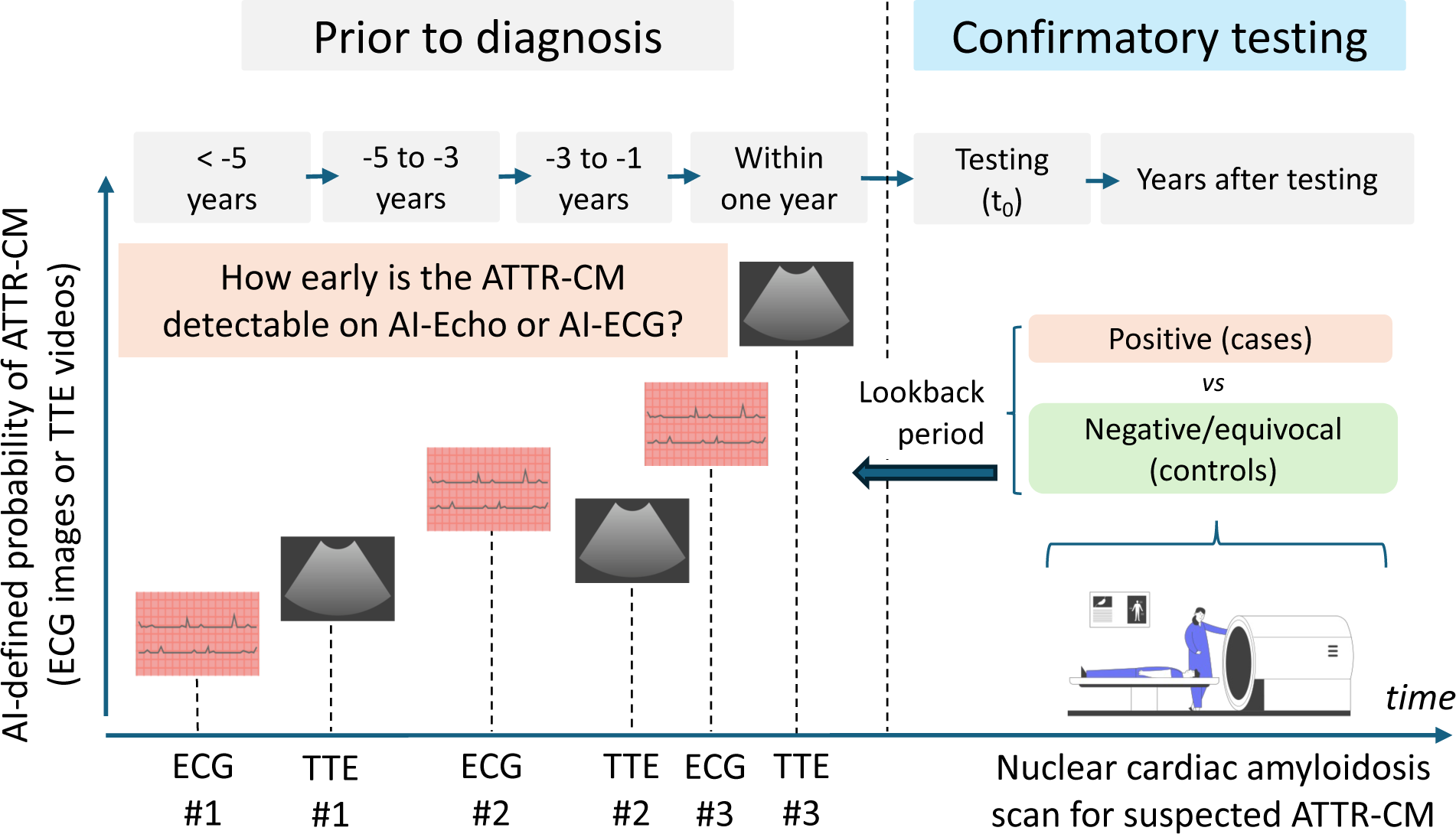
Study Population. Deep learning algorithms were trained to discriminate nuclear cardiac amyloid imaging-positive cases of ATTR-CM from age- and sex-matched controls using standard TTE videos or ECG images. These were subsequently deployed across independent sets of patients with longitudinal monitoring by TTE or ECG pre-dating their referral for nuclear cardiac amyloid testing. The overall objective was to examine the ability of the AI models to detect changes in TTE or ECG signatures that precede clinical disease and diagnosis. Such AI-enabled TTE or ECG signatures may be used to forecast the development of ATTR-CM, thus offering a standardized and scalable platform for longitudinal monitoring and screening in the community. AI: artificial intelligence; ATTR-CM: transthyretin amyloid cardiomyopathy; ECG: electrocardiography; TTE: transthoracic echocardiography.

## METHODS

### Study overview and data source

This was a retrospective study across two large and diverse health systems, namely the Yale-New Haven Health System (YNHHS, internal cohort), spanning 5 hospitals and affiliated clinic sites across Connecticut and Rhode Island, and the geographically distinct Houston Methodist Hospitals (HMH, external cohort, with 8 hospitals and affiliated clinic sites) in Houston, Texas. We leveraged existing AI-Echo and AI-ECG algorithmic pipelines and models previously designed to discriminate cases of ATTR-CM from age/sex-matched controls in YNHHS.^15–18^ This allowed us to extract the distinct echocardiographic and ECG signature of the ATTR-CM phenotype independently from each modality. Next, we deployed modality-specific models across all serial TTEs and ECGs performed in independent sets of adult individuals referred for nuclear cardiac amyloid testing in YNHHS (internal cohort) and HMH (external cohort). This design allowed us to assess the temporal evolution of AI-Echo and AI-ECG probabilities and their utility in discriminating future disease in the years before nuclear cardiac amyloid testing (**Figure 2**).^19^ Respective Institutional Review Boards approved the study protocol and waived the need for informed consent as the study involves secondary analysis of pre-existing data.

**Figure 2.**
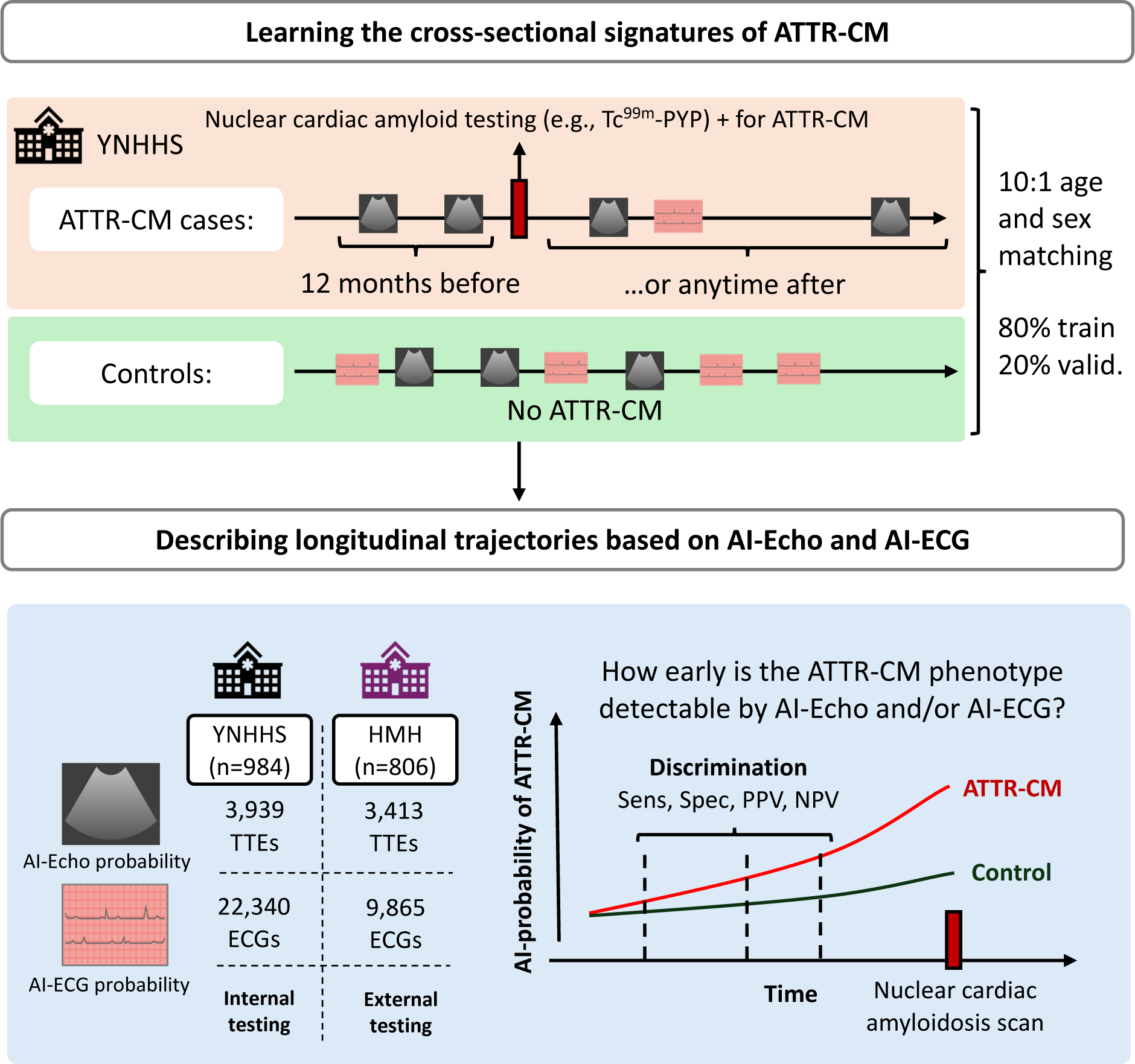
Study Design. AI-models were trained on transthoracic echocardiograms (TTE) and 12-lead electrocardiographic (ECG) images from patients with ATTR-CM (based on a positive nuclear cardiac amyloid test performed no more than 12 months after the index study) as well as age- and sex-matched controls without known disease across the Yale-New Haven Health System (YNHHS). Models were subsequently deployed across independent sets of patients in YNHHS, as well as an external set of patients from Houston Methodist Hospitals (HMH) who had sequential TTE or ECG performed in the years leading up to their referral for nuclear cardiac amyloid imaging. This design allowed us to evaluate the progression of AI-Echo or AI-ECG probabilities as non-invasive markers of pre-clinical ATTR-CM progression. AI: artificial intelligence; ATTR-CM: transthyretin amyloid cardiomyopathy; ECG: electrocardiography; HMH: Houston Methodist Hospitals; Tc^99m^-PYP: pyrophosphate; TTE: transthoracic echocardiography; YNHHS: Yale-New Haven Health System.

### Study definitions

As per our previous work,^15,16^ to define the AI-Echo and AI-ECG classifier for the cross-sectional detection of ATTR-CM, we identified any TTE or ECG studies performed up to one year before or any time after an abnormal nuclear cardiac amyloid study as positive for the ATTR-CM phenotype. This definition ensured the specificity of the label for ATTR-CM and was based on the median delay (latency) in ATTR-CM diagnosis that has been estimated at 12-13 months across contemporary studies.^13,20^ Across cohorts, nuclear cardiac amyloid testing was performed in accordance with the recommendations of the American Society of Nuclear Cardiology using single photon emission tomography-computed tomography [SPECT-CT] imaging with Tc^99m^-pyrophosphate [PYP] or other approved radiotracer. The final diagnosis of a positive study was adjudicated by the interpreting physician based on semi-quantitative visual grading (grade ≥2), or a heart-to-contralateral lung ratio of >1.5, where applicable.^19^ Clinical risk factors, co-morbidities and amyloidosis-related codes were extract by querying the respective EHR up to 12 months after the nuclear cardiac amyloidosis study (**Table S1**).

### Development cohorts: learning cross-sectional signatures of ATTR-CM

#### AI-Echo model development

##### AI-Echo training population

For the purposes of this study, we used a previously validated pipeline for training TTE-based classification algorithms,^16–18,21^ with training performed *de novo* to avoid overlap between individuals with longitudinal progression data and those included in training. Given the lower counts of echocardiograms relative to ECG studies and to ensure that patients with sequential studies were not seen during model training, participants who had ≥2 TTE studies before a nuclear cardiac amyloid exam were not included in the training or validation of the amyloid detection models but were reserved for the progression analysis. The diagnostic model development population included 308 studies from 101 unique individuals, split at a patient level into training and validation sets (80%, 20%) to develop an AI-Echo model for detecting concurrent ATTR-CM. Age- and sex-matched controls (10 controls per case) were sampled from individuals from the same period without a history of abnormal nuclear cardiac amyloid imaging or an ICD amyloidosis code specific for transthyretin amyloidosis (“E85.82”). The AI-Echo model development population is summarized in **Table S2**.

##### AI-Echo model training

We followed our previously described end-to-end pre-processing pipeline for echocardiographic studies stored in DICOM (Digital Imaging and Communications in Medicine) format, which involves deidentification, automated view classification steps, as well as standard augmentation by padding, random rotation, and horizontal flipping.^16,18^ We used a 3D ResNet-18 backbone, class-balanced loss function (weighted binary cross-entropy), the Adam optimizer, a learning rate of 10^-4^, a batch size of 56, a random dropout of 0.25, and label smoothing (α of 0.1), and trained our algorithm for a maximum of 30 epochs with patience (early stopping) set at 5 epochs. For predictive estimates for the full echocardiographic study, we used key echocardiographic views that included the left ventricle and left atrium (parasternal long axis, and any of the standard apical four-, three- or two-chamber views) as inputs and performed mean averaging of the output probabilities across these views. The full process is described in the **Supplement**.

#### AI-ECG model development

##### AI-ECG training population

The AI-ECG image model was trained in line with our previously described approach,^22–24^ and is reviewed in detail elsewhere.^15^ To increase the accuracy of our labels, we required that all controls had a TTE performed ≤15 days of the ECG during training but did not receive a diagnosis of ATTR-CM during follow-up. The AI-ECG development population is summarized in **Table S3**.

##### AI-ECG model training

Images of ECGs were generated from 12-lead recordings at a frequency of 500 Hz for 10 seconds collected on various machines (i.e., Philips PageWriter machines and GE MAC machines). We followed our previously described approach of standard transformation, calibration, plotting across various lead layout formats, baseline wander correction, and random augmentation.^22^ We used an EfficientNet-B3 backbone that was initialized using weights from a self-supervised biometric contrastive learning approach that we have previously defined.^23^ We used a class-balanced binary cross-entropy loss function, an Adam optimizer, gradient clipping, a learning rate of 64, a batch size of 10^-5^. The full process and training population is described in the **Supplemen**t, and separately.^15^

### Testing cohorts: evaluating pre-clinical ATTR-CM progression by AI-Echo & AI-ECG

The trained models were independently deployed across two geographically distinct patient populations drawn from YNHHS (September 2016 through January 2024: *internal cohort*) and HMH (March 2016 through May 2024: *external cohort*). The presence of two geographically distinct cohorts enabled a more robust assessment of performance across institutions and populations with different baseline risk and referral patterns. For the ECG analysis at YNHHS, we removed samples that had previously been used during the model’s training. We stratified both the YNHHS and HMH cohorts based on the results of the nuclear cardiac amyloid test as positive (first positive study for any individuals eventually diagnosed with ATTR-CM) or negative (first negative study, with no established diagnosis by the end of follow-up). Negative nuclear cardiac amyloid studies represented the controls since clinical suspicion was high enough to prompt referral for dedicated nuclear imaging. We directly deployed the AI-Echo and AI-ECG models to all TTE videos and ECG images from these participants (done before or after their nuclear cardiac amyloid test). Across cohorts, all TTE studies were available in standard DICOM format. In YNHHS (internal cohort), ECG images were available in a standard format as .png files, whereas in HMH (external cohort), these were exported directly as flattened .pdf files. To ensure transparency during testing in an external population, we embedded both the AI-ECG and AI-Echo models into executable applications that contained standardized environments and enabled direct inference on the TTE and ECG studies at HMH. The software applications can be made available upon reasonable request as part of a research collaboration. Interactive applications for both algorithms are openly available on our website for research use with deidentified data (see data availability statement).

### Statistical analysis

Continuous variables are presented as median [25^th^-75^th^ percentile] and compared using the Mann-Whitney test across two groups. Categorical variables are summarized as counts (and percentages) and compared across distinct groups using the χ^2^ test. During training, we summarized the discrimination performance of the AI-Echo and AI-ECG models using the area under the receiver operating characteristic curve [AUROC] for ATTR-CM with corresponding 95% confidence intervals (CI) derived from bootstrapping with 1,000 replications.

To assess the differential progression in AI-Echo and AI-ECG output probabilities (0 to 1) for the ATTR-CM phenotype across nuclear cardiac amyloid testing-positive vs negative participants, we fit a mixed-effects linear regression model with the AI-Echo or AI-ECG probability of ATTR-CM as the dependent variable, and the following fixed effects: nuclear cardiac amyloid imaging status (positive versus negative), the time difference between each ECG/TTE study, their interaction term, the time of nuclear cardiac amyloid imaging, age at the time of nuclear cardiac amyloid imaging, and sex. Given the correlatedness of observations within the same subject, participant was included as a random effect. We also present a similar analysis adjusting for echocardiographic parameters (left ventricular ejection fraction [LVEF], aortic stenosis presence and severity [none, mild, moderate, severe], left posterior wall thickness [LVPWd] and interventricular septum thickness at end-diastole [IVSd]), as extracted directly from the final echocardiography report. Furthermore, we derived annualized progression rates in the dependent variable (AI probability of ATTR-CM) across cases and controls by extracting the coefficients (and respective standard errors) for time and its interaction with nuclear cardiac amyloid imaging status from the previously fitted mixed linear model. Summary statistics (means and 95% confidence interval of mean) were also estimated and summarized across discrete time intervals (more than 5 years before nuclear cardiac amyloid imaging, 3 to 5 years before, 1 to 3 years before, last 12 months, or any time after). Across all participant-level analyses, if two or more tests of the same modality were performed in a given time window, we used the median of their individual probabilities.

Finally, we examined the ability of AI-Echo and AI-ECG to independently or jointly discriminate future ATTR-CM across these distinct time intervals. We present patient-level discrimination metrics (sensitivity, specificity, positive, negative predictive value) at a fixed threshold of 0.05 (but also across representative thresholds of 0.015, 0.10, 0.25 and/or 0.50) for various approaches: i) AI-Echo alone, ii) AI-ECG alone, and iii) joint testing among those who underwent both TTE and ECG in a given time window, using both an “Either” approach (≥1 required to be positive, vs all else), and a “Both” approach (both tests required to be positive, vs all else). For joint modelling, we excluded individuals who had missing data for one or more modalities in that window. This provided valid performance statistics based on temporally linked multimodal data. We provide 95% confidence intervals calculated by non-parametric bootstrapping with 1,000 replications. All statistical tests were two-sided with a significance level of 0.05 unless specified otherwise. All analyses were performed using Python version 3.11.2 (Python Software Foundation) and R version 4.2.3 (R Foundation).

## RESULTS

### Cross-sectional discrimination of ATTR-CM by AI-ECG and AI-Echo

We first confirmed the ability of AI-Echo and AI-ECG models to discriminate ATTR-CM. In the TTE held-out testing cohort that included 138 TTE cases and 1380 TTE control studies (median age 79 [IQR: 75, 84] years, 1166 [76.8%] male [**Table S2**]), the AI-Echo model reached a study-level AUROC of 0.93 (95%CI: 0.90-0.96). Similarly, in the ECG held-out testing cohort from YNHHS that included 139 ECG cases and 1390 ECG controls (median age 80 [IQR: 75, 86] years, 1,044 [68.3%] male [**Table S3**]) the AI-ECG model successfully discriminated ATTR-CM cases from controls with an AUROC of 0.91 (95%CI: 0.88-0.93).

### Pre-clinical cohort analysis

The validated AI-Echo and AI-ECG models were independently deployed across the pre-clinical progression cohorts. There were 3,939 unique TTEs and 22,340 ECGs in 984 unique individuals in the progression cohort at YNHHS, with 112 of these individuals (11.4%) having abnormal findings compatible with ATTR-CM on nuclear cardiac amyloid testing. At the external site, HMH, there were 3,413 TTEs and 9,865 ECGs in 806 participants, with 174 (21.6%) eventually diagnosed with ATTR-CM. Compared with negative cases, positive cases were older at the time of nuclear cardiac amyloid imaging (YNHHS: 82 [IQR 75, 86] vs 73 [IQR 64, 80] years, and HMH: 77 [IQR 70, 82] vs 67 [IQR 57, 75] years, all *p*<0.001) and more frequently men (YNHHS: n=77 [68.8%] vs n=471 [54.0%], and HMH: n=145 [83.3%] vs n=383 [60.6%], *p*<0.001). Further differences in representative comorbidities across groups are summarized in Table 1.

### Longitudinal phenotyping of the pre-clinical ATTR-CM phenotype by AI-Echo & AI-ECG

In the five years before nuclear cardiac amyloid testing was performed we observed a significant rise in the AI-derived probabilities of ATTR-CM among those who went on to test positive by nuclear cardiac amyloid imaging compared with their counterparts, findings that were consistent across cohorts (YNHHS: internal cohort; HMH: external cohort) and modalities (TTE and ECG) (**Figure 3**). Despite relative overlap in the AI probabilities >5 years before diagnosis between cases and controls, longitudinal follow-up demonstrated that the AI phenotypes started diverging as early as 3 years before diagnosis. In age- and sex-adjusted analyses in YNHHS, the annualized progression rate in AI-Echo probabilities was estimated at 2.6%/year [95%CI: 2.0%-3.1%/year] in cases vs 0.7%/year [95%CI: 0.5%-0.9%/year] in controls, and for AI-ECG at 1.4%/year [95%CI: 1.2%-1.7%/year] in cases vs 0.7%/year [95%CI: 0.6%-0.9%/year] in controls. These findings were replicated in the HMH cohort, with annualized progression rates of 1.8%/year [95%CI: 1.3%-2.3%] vs 0.3%/year [95%CI: 0.1%-0.5%] for AI-Echo, and 1.8%/year [95%CI: 1.2%-2.4%] vs 1.0% [95%CI: 0.7%-1.3%] for AI-ECG. Across all analyses, we observed a significant interaction when modelling nuclear cardiac amyloid testing positivity and time against AI-Echo and AI-ECG probabilities, thus supporting disproportionately faster progression rates among nuclear imaging-positive vs-negative cases in the years before diagnosis (**Table 2**).

**Figure 3.**
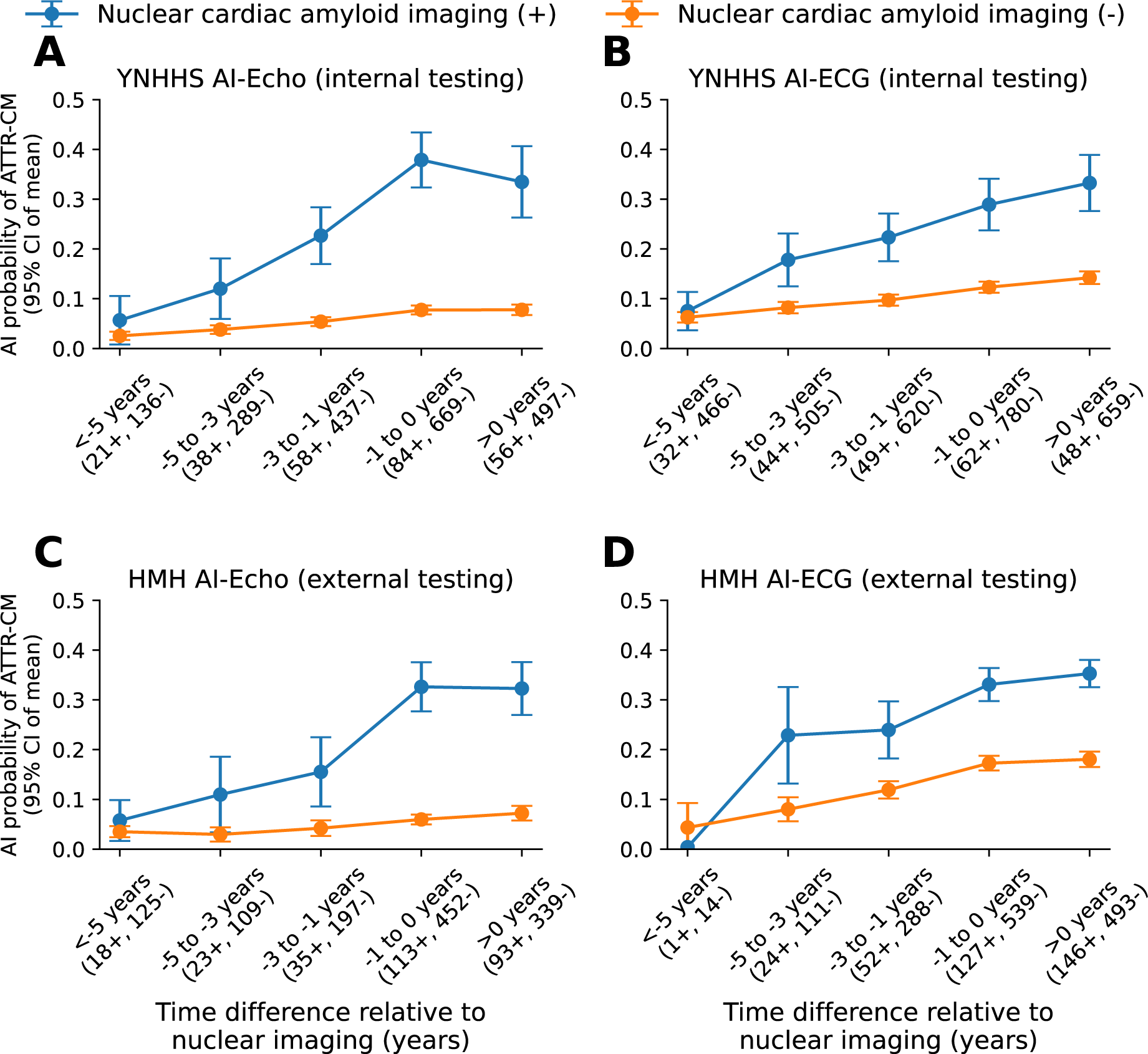
Longitudinal patient-level changes in AI-Echo and AI-ECG ATTR-CM probabilities across cohorts based on nuclear cardiac amyloid positivity. The panels illustrate the mean (with error bars denoting the 95% confidence interval of mean) of the AI-Echo and AI-ECG-derived probabilities across patients who went on to have a positive (blue color) vs negative (orange color) nuclear cardiac amyloid test across YNHHS (**A, B**) and HMH (**C, D**), respectively. The x axis denotes the time between the TTE/ECG and the timing of nuclear cardiac amyloid testing summarized across discrete time groups (negative time differences suggest that the TTE/ECG was performed before the nuclear cardiology exam). The brackets below each period along the x axis denote the number of unique positive and negative individuals in each period. If more than one study was available in a given period, we derived the median of all predictions in that timeframe. ATTR-CM: transthyretin amyloid cardiomyopathy; ECG: electrocardiography; HMH: Houston Methodist Hospitals; PYP: Tc^99m^-pyrophosphate; TTE: transthoracic echocardiography; YNHHS: Yale-New Haven Health System.

In further analyses, the echocardiographic findings were robust to multivariable adjustment for the presence and severity of aortic stenosis, LVEF, as well as LVPWd and IVSd, with annualized AI-Echo progression rates of 2.2% [95%CI: 1.7%-2.8%] among cases vs 0.6% [95%CI: 0.4%-0.8%] among controls in YNHHS (p_(group x time) interaction_<0.001). Finally, subgroup analyses of confirmed cases of AL (light chain) amyloidosis (n=19 [1.9%] individuals in YNHHS, and n=37 [4.6%] in HMH, based on diagnosis codes) suggested a similar rise in AI probabilities in the year before nuclear cardiac amyloid testing both in YNHHS and HMH (**Figure S1**).

### Diagnostic performance of AI-Echo and AI-ECG risk stratification for early ATTR-CM

Based on these observations, we evaluated the diagnostic performance of AI-Echo and AI-ECG, both as stand-alone tests and in combination, for discriminating ATTR-CM cases from controls across discrete time windows in the years preceding clinical diagnosis. **Figures 4-5** and **Tables S4-S5** summarize discrimination metrics across different strategies (AI-Echo alone, AI-ECG alone, “Either” [at least one positive, vs all else], or “Both” [double-positive, vs all else]), distinct pre-clinical time periods (3-5 years, 1-3 year, and up to 1 year before the nuclear cardiac amyloid exam), and AI-thresholds (0.015, 0.05, 0.10, 0.25, 0.50). For instance, 1-to-3 years before nuclear cardiac amyloid testing, AI-Echo values ≥0.05 had 78% and 49% sensitivity for discriminating subsequent ATTR-CM (with 69% and 79% specificity), whereas AI-ECG values ≥0.05 had 75% and 65% sensitivity (with 56% and 47% specificity) across YNHHS and HMH, respectively (**Figure 4** & **Table S4**). However, when considering joint testing among individuals with both AI-TTE and AI-ECG predictions 1-to-3 years before nuclear cardiac amyloid testing (n=433 [YNHHS] and 174 [HMH]), a double-negative screen (seen in 164 [37.9%] and 66 [37.9%] participants, vs all else) had 90.9% and 85.7% sensitivity (specificity of 40.3% and 41.2%), whereas a double-positive screen (seen in 78 [18.0%] and 26 [14.9%] participants, vs all else) had 85.5% and 88.9% specificity (sensitivity of 60.6% and 42.9%) across YNHHS and HMH, respectively (**Figure 5** & **Table S5**).

**Figure 4.**
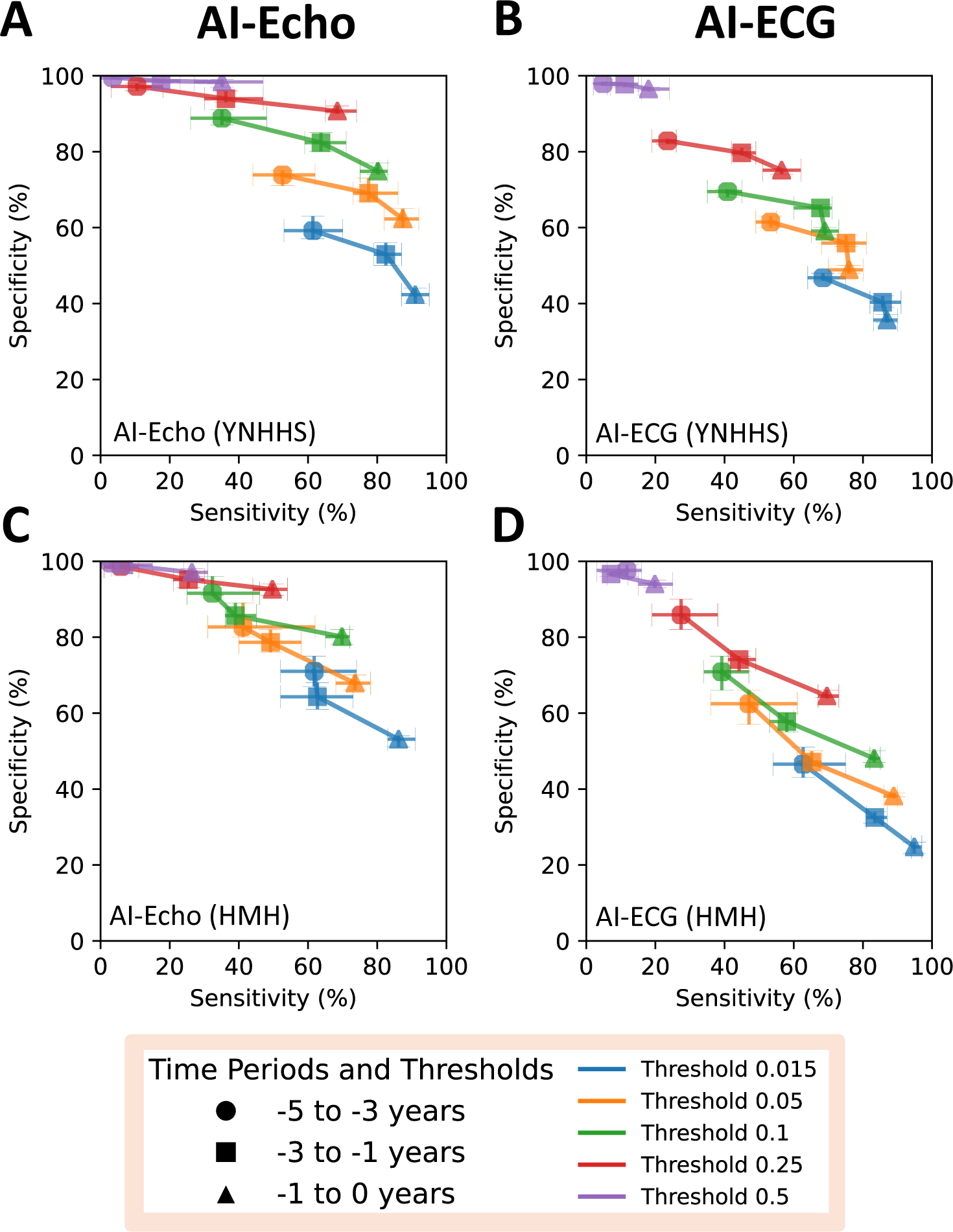
Sensitivity and specificity of AI-Echo and AI-ECG for ATTR-CM across pre-clinical stages. Each plot represents the sensitivity and specificity at a given threshold (0.015: blue, 0.05: orange, 0.1: green, 0.25: red, 0.5: purple) and a given time point (circle: 5 to 3 years; rectangle: 3 to 1 year(s) before; and triangle: year before nuclear cardiac amyloid testing across YNHHS (A-C) and HMH (D-F). The panels depict the evolution in predictions for AI-Echo (**A&C**), and a AI-ECG (**B&D**). The error bars in either direction denote the 95% confidence intervals for sensitivity (x axis), or specificity (y axis) derived from bootstrapping with 1000 replications. ATTR-CM: transthyretin amyloid cardiomyopathy; ECG: electrocardiography; HMH: Houston Methodist Hospitals; TTE: transthoracic echocardiography; YNHHS: Yale-New Haven Health System.

**Figure 5.**
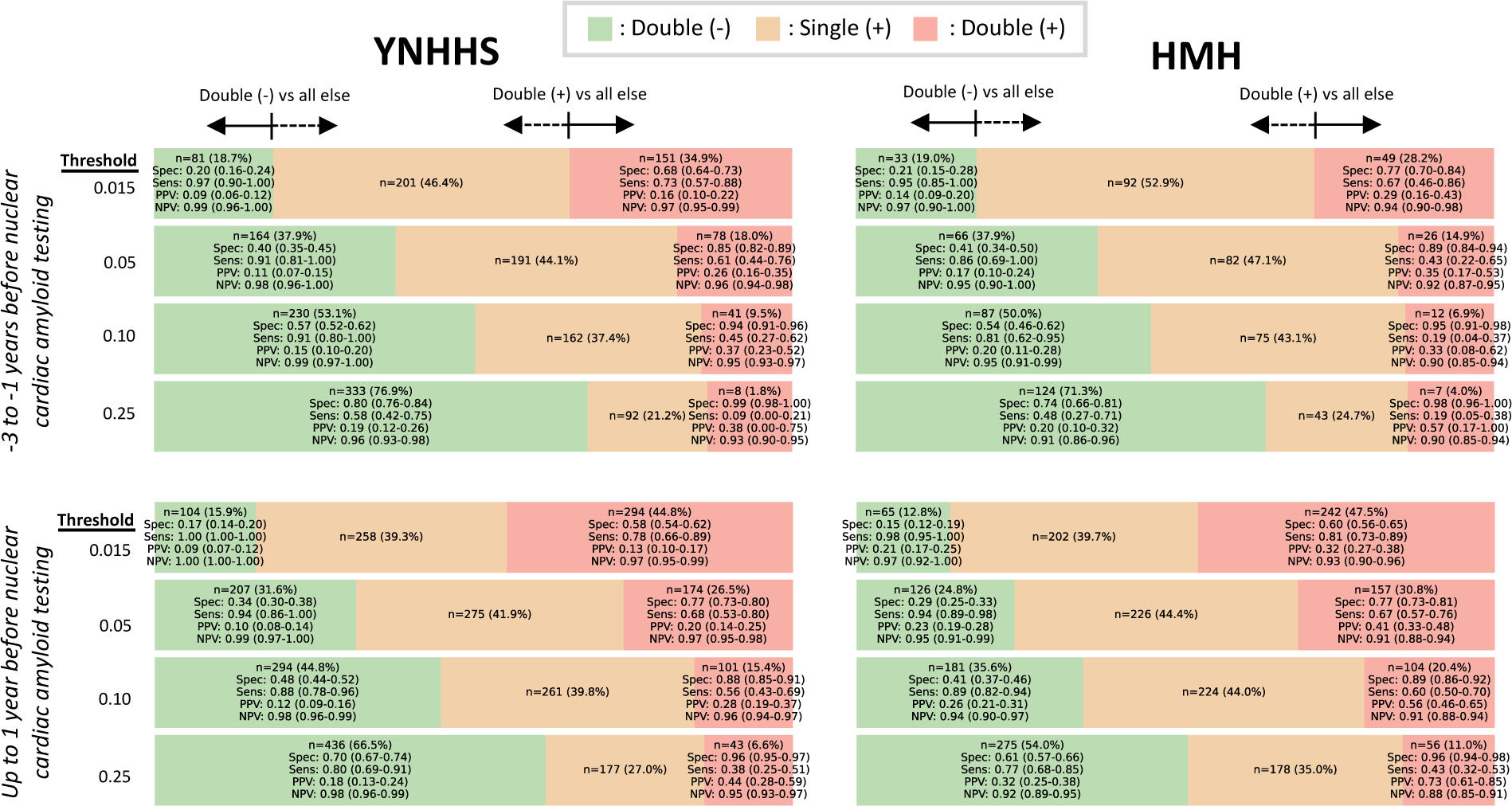
Discrimination performance of joint AI-Echo and AI-ECG testing for future ATTR-CM across pre-clinical stages. We present the observed counts, and discrimination (sensitivity, specificity, positive [PPV] and negative predictive value [NPV]) for joint testing by both AI-Echo and AI-ECG across thresholds and timepoints. We present two operating points, a sensitive one where we compare double-negatives versus everyone else, and a more specific one where we compare double-positives against everyone else. We also present 95% confidence intervals derived from bootstrapping with 1000 replications. AI: artificial intelligence; ATTR-CM: transthyretin amyloid cardiomyopathy; ECG: electrocardiography; HMH: Houston Methodist Hospitals; TTE: transthoracic echocardiography; YNHHS: Yale-New Haven Health System.

## DISCUSSION

In two large, diverse, and geographically distinct health system-based cohorts, we demonstrate that AI applied to standard TTE videos and ECG images enables scalable risk stratification and screening for ATTR-CM early in its course and before clinical diagnosis through standard pathways. Our findings introduce a new paradigm in which AI-enhanced interpretation of accessible diagnostic tests may be used to detect ATTR-associated changes which precede the clinical diagnosis of ATTR-CM by several years (**Graphical Abstract**). We provide evidence to support the generalizability of these observations across distinct cohorts and further illustrate their dynamic nature in line with the natural history of ATTR-CM progression. These findings may support the use of AI-Echo and AI-ECG in identifying at-risk individuals, guiding downstream diagnostics and informing the application of novel risk-modifying therapies while also enabling monitoring through cost-effective technologies.

Our findings should be interpreted in the context of recent evidence on the changing epidemiological and therapeutic landscape of ATTR-CM. Given advances in the non-invasive diagnosis by nuclear cardiac amyloid imaging and cardiac magnetic resonance (CMR) imaging, referrals to amyloidosis centers have been increasing.^13^ While a U.K.-based study estimated that the median duration of symptoms prior to diagnosis has decreased from 36 months in 2002 to 12 months in 2021,^13^ multinational registries demonstrate persistent delays from symptom onset to ATTR-CM diagnosis that often exceed two years.^25^ There is also a growing recognition that myocardial ATTR deposition often co-exists with prevalent conditions, such as aortic stenosis and heart failure with preserved ejection fraction,^2–4^ thus highlighting an emerging need for scalable and cost-efficient screening tool that can be deployed serially in at-risk populations.

To date, efforts to develop reliable prognostic biomarkers for ATTR-CM have been hampered by the low prevalence in the community, complex etiological and pathophysiological profile, and substantial heterogeneity across cases.^1^ However, rapid advances in the therapeutic landscape of ATTR-CM have revealed a gap in scalable diagnostics to monitor the pre-clinical stages of the condition. Most prognostic markers have been evaluated among patients with an existing diagnosis, such as NT-proBNP (N-terminal pro-Brain Natriuretic Peptide) levels or outpatient intensification of diuretics, which consistently portend worse prognosis among patients with ATTR-CM.^26^ Furthermore, while nuclear cardiac amyloid imaging represents an excellent non-invasive alternative to traditional biopsy,^27,28^ it is expensive, not widely available, and associated with radiation exposure, all features that preclude its use in longitudinal monitoring. Recently, attention has shifted to maximizing inference from easily scalable modalities performed during initial patient evaluation. AI methods directly applied to ECG signals and TTE videos have shown potential in detecting distinct electrical and structural signatures associated with the cardiac amyloidosis phenotype.^29–31^ However, these models have been limited to individuals with established diagnosis and do not inform the use of these models in the risk stratification of preclinical ATTR-CM. Thus, our study introduces a novel role for AI-ECG and AI-Echo in the screening, risk stratification, and longitudinal disease monitoring of pre-clinical ATTR-CM through accessible and scalable biomarkers. Expanding AI-assisted phenotyping to the pre-clinical stages of ATTR-CM represents a critical unmet need to boost its timely recognition, monitor its progression, and eventually guide the deployment of disease-modifying therapies.^32^

As the potential eligibility pool for new therapies expands further, AI-enabled phenotyping could also guide optimal case selection during this pre-clinical stage and identify individuals who may derive the greatest benefit from early intervention based on an objectively quantifiable and dynamically evolving phenotype. Moreover, since many variant forms (i.e., V122I/pV142I) are more prevalent among traditionally disadvantaged communities, including racial and ethnic monitories (i.e., individuals of African or Hispanic/Latino ancestry), AI-enabled interpretation of accessible diagnostic modalities may also improve access to timely diagnostic care,^33–35^ given that they represent the most commonly accessible form of ECGs available to the end-users. The feasibility, scalability, and performance of this approach and the models covered in this manuscript are currently explored within multi-center federated efforts such as the TRACE-AI network study which are designed to evaluate the burden of undiagnosed ATTR-CM across health systems and investigate strategies to minimize diagnostic delays. With the arrival of a rapidly expanding armamentarium of therapeutic agents,^1,7,8,10–12,36^ AI-Echo and AI-ECG-enabled risk stratification methods may identify at-risk individuals, guide targeted enrichment in prospective clinical trials, and further expand our knowledge on the pre-clinical stages of the disease, its trajectory, and the ability to modify it. This concept of early preventive intervention is already being explored in phase 3 trials of transthyretin stabilizers in individuals with pathogenic *TTR* variants, however assessment of response requires access to gold-standard nuclear cardiac amyloid imaging.^37^

Ultimately, our method may be applied both retrospectively and prospectively across repositories of cardiac diagnostics, as well as at the point of care, to identify, track, and risk stratify the earliest pre-clinical stages of ATTR-CM from accessible modalities. A sample implementation framework is summarized in **Figure S3**, although this would require prospective validation before integration into clinical pathways. Here, a double negative screen demonstrates a high NPV, potentially deferring the need for further testing, whereas a double-positive screen exhibits high specificity and PPV, enabling early identification of at-risk patients before clinical suspicion arises or nuclear cardiac amyloid testing is performed. Although the proposed strategy relies on single, one-time assessment, longitudinal monitoring with AI-Echo and AI-ECG could provide additional insights in equivocal cases or when clinical suspicion persists. By enabling longitudinal monitoring through standardized interpretation of scalable diagnostic modalities, our proposed strategy also enhances precision by minimizing rater variability and mitigating potential sources of referral and diagnostic bias associated with limited access to advanced imaging facilities.^38,39^

### Limitations

Certain limitations merit consideration. First, this was a retrospective analysis of individuals in whom clinical suspicion for ATTR-CM prompted a referral for dedicated nuclear cardiac amyloid testing. Similarly, ECG and TTE studies performed before or after clinical diagnosis were part of clinical care, thus introducing selection bias. As a result, it is unclear whether these findings can generalize to undiagnosed patients with pre-clinical ATTR-CM in the community who were never screened by ECG, TTE, or nuclear cardiac amyloid testing. The joint testing analysis also reflects a non-randomly selected subset of individuals who had both tests done in the same period, which may affect the generalizability of these observations. On this note, while this method shows promise for early disease detection, its potential must be balanced against the risk of false discovery and unnecessary downstream testing, both of which require rigorous evaluation in prospective clinical trials. Second, nuclear imaging-negative cases were younger and less likely to be male than their nuclear imaging-positive counterparts.

Although the longitudinal trends were independent of age and sex, a negative study does not rule out the future development of the disease, particularly if testing was done at very early stages. Third, we relied on nuclear cardiac amyloid imaging with bone-avid radiotracers such as Tc^99m^-PYP which are known to be highly sensitive and specific for ATTR-CM using established quantitative and semi-quantitative thresholds.^28^ However, since these studies were performed as part of clinical care, there was no consistent multimodal assessment, or histological confirmation. Similarly, there was no consistent genotyping to assess differential trends across variant and wild-type forms of the disease. Fourth, our case definition allowed for some overlap forms of nuclear imaging-positive AL amyloidosis (possible AL/ATTR overlap) to be included in the cases and nuclear imaging-negative forms of non-ATTR (e.g. AL amyloidosis) to be included in the controls. This reflects the limitations of non-invasive imaging and the challenges of diagnosing cardiac amyloidosis. Nevertheless, subgroup analyses suggest that a similar, possibly steeper rise may emerge for individuals with AL amyloidosis, but this requires further investigation in dedicated studies. Fifth, ATTR-CM is a dynamic process, and a negative screen does not rule out future disease which may manifest several years later. In these cases, longitudinal monitoring of AI-derived probabilities may offer an observer-independent, quantitative metric to examine longitudinal changes. For screening purposes, however, decisions should likely rely on the most recent exam, rather than retrospective, historical trends. Sixth, care practices may differ across centers, and the participants included in our analysis may have been evaluated by different teams of general practitioners, cardiologists, or cardiomyopathy specialists at different parts of their care, thus introducing potential heterogeneity in the cohorts. We also observed differences in demographics and the prevalence of abnormal nuclear cardiac amyloid test results across the internal and external cohorts, although this did not affect the generalizability of our observations. Ultimately, the broader paradigm suggested by this analysis requires further evaluation in prospective clinical cohorts or trials with protocolized imaging of at-risk populations.

## CONCLUSIONS

AI technology applied directly to echocardiography and ECG images may enable scalable identification and risk stratification of pre-clinical ATTR-CM. These findings suggest a possible role for AI-enabled interpretation of routinely performed cardiac investigations to flag individuals at high risk of progressing to clinical ATTR-CM.

## DATA AVAILABILITY STATEMENT

The underlying data represent protected health information. To protect patient privacy, the local Institutional Review Boards within each center do not allow the sharing of these data. Browser-accessible interactive versions of the AI-Echo and AI-ECG models are openly available on our Lab’s website for research use (AI-ECG: https://www.cards-lab.org/ecgvision-attrcm and AI-Echo: https://www.cards-lab.org/aiecho-attrcm). Executable applications of the AI-ECG and AI-TTE models can be made available for research use by contacting the corresponding author.

