## Supplemental Material for "Tracking the Preclinical Progression of Transthyretin Amyloid Cardiomyopathy Using Artificial Intelligence-Enabled Electrocardiography and Echocardiography"

Evangelos K. Oikonomou MD DPhil[a,b], Veer Sangha BS[b,c], Sumukh Vasisht Shankar MS[a,b], Andreas Coppi PhD[b,d], Harlan M. Krumholz MD SM[a,d], Khurram Nasir MD MPH,[e,f,g] Edward J. Miller MD PhD[a], Cesia Gallegos-Kattan MD MHS[a], Mouaz H. Al-Mallah MD MSc[g], Sadeer Al-Kindi MD[g,h], Rohan Khera MD MS[a,b,d,i,j*]

[a] Section of Cardiovascular Medicine, Department of Internal Medicine, Yale School of Medicine, New Haven, CT, USA
[b] Cardiovascular Data Science (CarDS) Lab, Yale School of Medicine, New Haven, CT, USA
[c] Department of Engineering Science, University of Oxford, Oxford, UK
[d] Center for Outcomes Research and Evaluation, Yale-New Haven Hospital, New Haven, CT, USA
[e] Division of Cardiovascular Prevention and Wellness, Department of Cardiology, Houston Methodist DeBakey Heart & Vascular Center, Houston, TX, USA
[f] Houston Methodist-Rice Digital Health Institute, Houston, TX, USA
[g] Houston Methodist DeBakey Heart and Vascular Center, Houston, TX, USA
[h] Center for Cardiovascular Computational & Precision Health, Houston Methodist DeBakey Heart & Vascular Center, Houston, TX, USA
[i] Section of Biomedical Informatics and Data Science, Yale School of Medicine, New Haven, CT, USA
[j] Section of Health Informatics, Department of Biostatistics, Yale School of Public Health, New Haven, CT, USA

**Table of contents:**
**Supplemental Methods:** pages 2, 3
**Supplemental Tables S1-5:** pages 4-8
**Supplemental Figures S1-2:** pages 9-10
**Supplemental References:** page 11

**[*]Correspondence to:**
Rohan Khera, MD, MS
195 Church St, 6th Floor, New Haven, CT 06510
203-764-5885;

### Supplemental Methods

#### AI-Echo model training pipeline

***TTE pre-processing:*** We implemented our previously published end-to-end pre-processing pipeline on echocardiographic studies that studies stored in DICOM format, which involves loading the pixel data, masking out pixels in the periphery to remove identifying information and converting to Audio Video Interleave (.AVI) format.[1] First, we randomly sampled ten frames from each video, down-sampled to 224x224 pixels, and fed these frames through a previously validated VGG19 convolutional neural network (CNN) that enables video-level classification of 18 echocardiographic views by assigning a probability that a given video corresponds to a standard anatomical view (with probabilities adding up to 1 across all views).[2] A predicted view was then assigned based on the view class that has the highest probability. Next, we performed more thorough cleaning and de-identification by binarizing each video frame with a fixed threshold, masking out all pixels outside the convex hull of the largest contour, and down-sampling to 112x112 pixels, as described in our previous work.[1,3] We initialized a 3D-ResNet18 CNN architecture by using pre-trained weights from the Kinetics-400 dataset, and trained a binary, video-level classifier to detect the presence of ATTR-CM from controls. As part of data augmentation, we applied random zero padding by up to 8 pixels in each spatial dimension, random horizontal flipping with (probability 0.5), and a random rotation within -10 and 10 degrees (probability 0.5). After augmentation, each video clip's intensities were normalized to 0-1 and standardized using the channel-wise means and standard deviations from the Kinetics-400 training dataset.

***AI-Echo model training and inference:*** Training was performed across all videos included in each study, as a form of natural augmentation that prompts the model to learn from variable views. Models were trained on four NVIDIA Tesla T4 GPUs with a class-balanced loss function (weighted binary cross-entropy), the Adam optimizer, a learning rate of $10^{-4}$, a batch size of 56 to maximize GPU utilization, and a random dropout of 0.25, and label smoothing ($\alpha$=0.1). We used class-balanced binary cross entropy loss and trained for a maximum of 30 epochs with patience (early stopping) set at 5 epochs. We randomly sampled video clips of 16 frames and sampling one out of every five frames to enable a global capture of the cardiac cycle. We applied optional padding with empty frames along the temporal axis if needed. At the time of inference, we averaged four 16-frame-clip-level predictions to obtain video-level predictions for each label. To provide study-level estimates, we restricted our analysis to key echocardiographic views that included the left ventricle and left atrium (parasternal long axis, and any of the standard apical four-, three- or two-chamber views) and performed mean averaging of the output probabilities.

#### AI-ECG model training pipeline

***AI-ECG model inference*:**

Baseline 12-lead ECG studies were extracted and pre-processed to create images of ECGs from standard 12-lead recordings at a frequency of 500 Hz for 10 seconds collected on various machines (i.e., Philips PageWriter machines and GE MAC machines).[4] ECG signals were transformed to ECG images with a calibration of 10 mm/mV, followed by conversion to grayscale and down-sampling to 300 x 300 pixels. Four formats of ECG images were plotted (standard, two-rhythm, alternate, and shuffled standard format), as described previously. Baseline wander correction was performed during the creation of these ECG images, aligned with preprocessing performed by ECG vendors prior to ECG printout creation. All images were rotated a random amount between -10 and 10 degrees before being input into the model. We used a class-balanced loss function (weighted binary cross-entropy), an Adam optimizer, gradient clipping, a batch size of 64, and a learning rate of $10^{-5}$, consistent with our previous AI-ECG studies. This approach simulates a clinical workflow where a provider may wish to directly access the algorithm through their browser or smartphone application by taking a photo or screenshot of a 12-lead ECG, as previously described.[4–6] The full AI-ECG model development study is described elsewhere.[7]

**Table S1 | ICD (International Classifications of Disease)-10 codes used.**

| Condition | ICD-10 codes |
|---|---|
| **Hypertension** | 'I10', 'I11', 'I110', 'I119', 'I12', 'I120', 'I129', 'I13', 'I130', 'I131', 'I132', 'I139', 'I674', 'O10', 'O100', 'O101', 'O102', 'O103', 'O109', 'O11' |
| **Diabetes Mellitus** | 'E10', 'E100', 'E101', 'E102', 'E103', 'E104', 'E105', 'E106', 'E107', 'E108', 'E109', 'E11', 'E110', 'E111', 'E112', 'E113', 'E114', 'E115', 'E116', 'E117', 'E118', 'E119', 'E12', 'E120', 'E121', 'E122', 'E123', 'E124', 'E125', 'E126', 'E127', 'E128', 'E129', 'E13', 'E130', 'E131', 'E132', 'E133', 'E134', 'E135', 'E136', 'E137', 'E138', 'E139', 'E14', 'E140', 'E141', 'E142', 'E143', 'E144', 'E145', 'E146', 'E147', 'E148', 'E149' |
| **Chronic Kidney Disease** | 'I12', 'I120', 'I13', 'I130', 'I131', 'I132', 'I139', 'N18', 'N180', 'N181', 'N182', 'N183', 'N184', 'N185', 'N188', 'N189', 'Z49', 'Z490', 'Z491', 'Z492' |
| **Heart Failure** | 'I110', 'I130', 'I132', 'I50', 'I500', 'I501', 'I509' |
| **Acute Myocardial Infarction** | 'I21', 'I22', 'I23', 'I240', 'I248', 'I249' |
| **Ischemic Heart Disease** | 'I20', 'I200', 'I208', 'I209', 'I21', 'I210', 'I211', 'I212', 'I213', 'I214', 'I219', 'I21X', 'I22', 'I220', 'I221', 'I228', 'I229', 'I23', 'I230', 'I231', 'I232', 'I233', 'I234', 'I235', 'I236', 'I238', 'I24', 'I240', 'I241', 'I248', 'I249', 'I25', 'I250', 'I251', 'I252', 'I255', 'I256', 'I258', 'I259', 'Z951', 'Z955', "Z9861", "Z951", "I2570", "I2571", "I2572", "I2573", "I2579", "I25810" |
| **Peripheral Arterial Disease** | 'I702', 'I7020', 'I7021', 'I742', 'I743', 'I744' |
| **Stroke** | 'G45', 'G450', 'G451', 'G452', 'G453', 'G454', 'G458', 'G459', 'I63', 'I630', 'I631', 'I632', 'I633', 'I634', 'I635', 'I638', 'I639', 'I64', 'I65', 'I650', 'I651', 'I652', 'I653', 'I658', 'I659', 'I66', 'I660', 'I661', 'I662', 'I663', 'I664', 'I668', 'I669', 'I672', 'I693', I694' |
| **Cardiomyopathy** | "I420", "I421", "I422", "I425", "I428", "I429", "I431", "I438" |
| **Hypertrophic cardiomyopathy** | "I421", "I422" |
| **Amyloidosis (any)** | “E85” |
| **Light Chain Amyloidosis** | “E85.81” |

**Table S2 | Study-level demographics for AI-Echo model development.**

| | | Training/Validation | | Testing | |
|---|---|---|---|---|---|
| | | **Case** | **Control** | **Case** | **Control** |
| **n** | | 308 | 3080 | 138 | 1380 |
| **Age (years)** | | 82 [77,86] | 81 [77,86] | 79 [75,84] | 79 [75,84] |
| **Gender** | Female | 69 (22.4) | 690 (22.4) | 32 (23.2) | 320 (23.2) |
| | Male | 239 (77.6) | 2390 (77.6) | 106 (76.8) | 1060 (76.8) |
| **Race** | Asian | - | 27 (0.9) | - | 13 (0.9) |
| | Black/African American | 50 (16.2) | 197 (6.4) | 43 (31.2) | 109 (7.9) |
| | Other | 3 (1.0) | 60 (1.9) | 3 (2.2) | 26 (1.9) |
| | Unknown | 5 (1.6) | 108 (3.5) | 4 (2.9) | 71 (5.1) |
| | White or Caucasian | 250 (81.2) | 2688 (87.3) | 88 (63.8) | 1161 (84.1) |
| **Ethnicity** | Hispanic or Latino | 9 (2.9) | 140 (4.5) | 15 (10.9) | 70 (5.1) |
| | Not Hispanic or Latino | 292 (94.8) | 2853 (92.6) | 123 (89.1) | 1254 (90.9) |
| | Unknown | 7 (2.3) | 87 (2.8) | - | 56 (4.1) |
| **Cardiovascular Risk factors** | | | | | |
| Hypertension | | 283 (91.9) | 2840 (92.2) | 123 (89.1) | 1236 (89.6) |
| Diabetes mellitus | | 99 (32.1) | 989 (32.1) | 48 (34.8) | 451 (32.7) |
| Chronic Kidney Disease | | 174 (56.5) | 1047 (34.0) | 67 (48.6) | 412 (29.9) |
| Acute Myocardial Infarction | | 25 (8.1) | 349 (11.3) | 18 (13.0) | 131 (9.5) |
| Ischemic Heart Disease | | 141 (45.8) | 1342 (43.6) | 56 (40.6) | 533 (38.6) |
| Peripheral Arterial Disease | | 9 (2.9) | 48 (1.6) | 2 (1.4) | 19 (1.4) |
| Stroke | | 51 (16.6) | 771 (25.0) | 43 (31.2) | 317 (23.0) |
| Heart Failure | | 279 (90.6) | 1574 (51.1) | 137 (99.3) | 642 (46.5) |
| Cardiomyopathy | | 163 (52.9) | 1021 (33.1) | 108 (78.3) | 485 (35.1) |
| Hypertrophic Cardiomyopathy (HCM) | | 34 (11.0) | 437 (14.2) | 15 (10.9) | 271 (19.6) |
| **Any amyloidosis ICD code** | | 305 (99.0) | 201 (6.5) | 138 (100.0) | 90 (6.5) |
| Non-neuropathic hereditary amyloidosis | | - | 3 (0.1) | 5 (3.6) | 6 (0.4) |
| Neuropathic hereditary amyloidosis | | 8 (2.6) | 5 (0.2) | 19 (13.8) | 3 (0.2) |
| Heredofamilial amyloidosis (unspecified) | | 8 (2.6) | 1 (0.0) | 12 (8.7) | 3 (0.2) |
| Secondary systemic amyloidosis | | 16 (5.2) | 18 (0.6) | 14 (10.1) | 6 (0.4) |
| Organ-limited amyloidosis | | 290 (94.2) | 158 (5.1) | 138 (100.0) | 75 (5.4) |
| Other amyloidosis | | 7 (2.3) | 16 (0.5) | 10 (7.2) | 10 (0.7) |
| Light-chain amyloidosis | | 4 (1.3) | 24 (0.8) | 7 (5.1) | 16 (1.2) |
| Wild-type TTR amyloidosis | | 69 (22.4) | 28 (0.9) | 66 (47.8) | 20 (1.4) |
| Amyloidosis, unspecified | | 166 (53.9) | 102 (3.3) | 103 (74.6) | 53 (3.8) |

Summary statistics are presented as counts (n) with valid percentages (%) or median [$25^{th}$-$75^{th}$ percentile].

### Table S3 | Study-level demographics for AI-ECG model development.

| | | Training/Validation | | Testing | |
|---|---|---|---|---|---|
| | | Case | Control | Case | Control |
| **n** | | 1011 | 10110 | 139 | 1390 |
| **Age (years)** | | 79 [70,85] | 77 [70,84] | 80 [75,86] | 80 [75,86] |
| **Gender** | Female | 180 (17.8) | 2414 (23.9) | 44 (31.7) | 441 (31.7) |
| | Male | 831 (82.2) | 7696 (76.1) | 95 (68.3) | 949 (68.3) |
| **Race** | Asian | - | 126 (1.2) | - | 12 (0.9) |
| | Black/African American | 176 (17.4) | 1065 (10.5) | 35 (25.2) | 93 (6.7) |
| | Other | 15 (1.5) | 349 (3.5) | - | 42 (3.0) |
| | Unknown | 50 (4.9) | 366 (3.6) | 3 (2.2) | 78 (5.6) |
| | White or Caucasian | 770 (76.2) | 8204 (81.1) | 101 (72.7) | 1165 (83.8) |
| **Ethnicity** | Hispanic or Latino | 58 (5.7) | 653 (6.5) | 4 (2.9) | 72 (5.2) |
| | Not Hispanic or Latino | 931 (92.1) | 9294 (91.9) | 131 (94.2) | 1240 (89.2) |
| | Unknown | 22 (2.2) | 163 (1.6) | 4 (2.9) | 78 (5.6) |
| **Cardiovascular Risk factors** | | | | | |
| Hypertension | | 876 (86.6) | 8651 (85.6) | 132 (95.0) | 1050 (75.5) |
| Diabetes mellitus | | 310 (30.7) | 3535 (35.0) | 30 (21.6) | 349 (25.1) |
| Chronic Kidney Disease | | 432 (42.7) | 3242 (32.1) | 49 (35.3) | 238 (17.1) |
| Acute Myocardial Infarction | | 102 (10.1) | 1419 (14.0) | 20 (14.4) | 45 (3.2) |
| Ischemic Heart Disease | | 410 (40.6) | 4626 (45.8) | 52 (37.4) | 296 (21.3) |
| Peripheral Arterial Disease | | 32 (3.2) | 211 (2.1) | | 6 (0.4) |
| Stroke | | 195 (19.3) | 2617 (25.9) | 23 (16.5) | 217 (15.6) |
| Heart Failure | | 895 (88.5) | 4457 (44.1) | 99 (71.2) | 262 (18.8) |
| Cardiomyopathy | | 650 (64.3) | 2356 (23.3) | 51 (36.7) | 121 (8.7) |
| Hypertrophic Cardiomyopathy (HCM) | | 138 (13.6) | 163 (1.6) | 7 (5.0) | 6 (0.4) |
| **Any amyloidosis ICD code** | | 959 (94.9) | 51 (0.5) | 81 (58.3) | 9 (0.6) |
| Non-neuropathic hereditary amyloidosis (E85.0) | | 15 (1.5) | 1 (0.0) | - | - |
| Neuropathic hereditary amyloidosis (E85.1) | | 60 (5.9) | - | - | - |
| Heredofamilial amyloidosis (unspecified) (E85.2) | | 45 (4.5) | - | - | 1 (0.1) |
| Secondary systemic amyloidosis (E85.3) | | 96 (9.5) | 8 (0.1) | - | 1 (0.1) |
| Organ-limited amyloidosis (E85.4) | | 921 (91.1) | 29 (0.3) | 81 (58.3) | 7 (0.5) |
| Other amyloidosis (E85.8) | | 55 (5.4) | 5 (0.0) | - | 3 (0.2) |
| Light-chain amyloidosis (E85.81) | | 15 (1.5) | 11 (0.1) | 6 (4.3) | 1 (0.1) |
| Wild-type transthyretin amyloidosis (E85.82) | | 346 (34.2) | 2 (0.0) | 13 (9.4) | - |
| Amyloidosis, unspecified (E85.9) | | 586 (58.0) | 25 (0.2) | 39 (28.1) | 4 (0.3) |

Summary statistics are presented as counts (n) with valid percentages (%) or median [$25^{th}$-$75^{th}$ percentile].

**Table S4 | Sensitivity & specificity across timepoints based on modality-specific testing.**

| Method | Time period | Threshold | Yale-New Haven Health System (YNHHS) Sensitivity (95% CI) | YNHHS Specificity (95% CI) | YNHHS PPV (95% CI) | YNHHS NPV (95% CI) | Houston Methodist Hospitals (HMH) Sensitivity (95% CI) | HMH Specificity (95% CI) | HMH PPV (95% CI) | HMH NPV (95% CI) |
|---|---|---|---|---|---|---|---|---|---|---|
| AI-ECG | -3 to -1 years | 0.015 | 0.86 (0.82-0.91) | 0.40 (0.39-0.42) | 0.07 (0.06-0.07) | 0.98 (0.98-0.99) | 0.83 (0.81-0.87) | 0.33 (0.31-0.34) | 0.15 (0.13-0.15) | 0.93 (0.92-0.95) |
| | -1 to 0 years | | 0.87 (0.83-0.90) | 0.36 (0.35-0.37) | 0.06 (0.06-0.07) | 0.98 (0.98-0.99) | 0.95 (0.94-0.97) | 0.25 (0.23-0.26) | 0.17 (0.16-0.19) | 0.97 (0.95-0.97) |
| | -3 to -1 years | 0.05 | 0.75 (0.68-0.81) | 0.56 (0.55-0.58) | 0.08 (0.07-0.09) | 0.98 (0.97-0.98) | 0.65 (0.61-0.68) | 0.47 (0.46-0.50) | 0.15 (0.13-0.17) | 0.91 (0.89-0.92) |
| | -1 to 0 years | | 0.76 (0.70-0.80) | 0.49 (0.48-0.50) | 0.07 (0.06-0.08) | 0.98 (0.97-0.98) | 0.89 (0.86-0.90) | 0.38 (0.38-0.39) | 0.19 (0.18-0.21) | 0.95 (0.94-0.96) |
| | -3 to -1 years | 0.1 | 0.68 (0.60-0.71) | 0.65 (0.65-0.66) | 0.09 (0.07-0.10) | 0.98 (0.97-0.98) | 0.58 (0.53-0.60) | 0.58 (0.55-0.60) | 0.16 (0.14-0.16) | 0.91 (0.89-0.92) |
| | -1 to 0 years | | 0.69 (0.65-0.73) | 0.59 (0.58-0.60) | 0.08 (0.06-0.09) | 0.97 (0.97-0.98) | 0.83 (0.82-0.85) | 0.48 (0.47-0.50) | 0.21 (0.20-0.22) | 0.95 (0.94-0.95) |
| | -3 to -1 years | 0.25 | 0.45 (0.42-0.49) | 0.80 (0.79-0.81) | 0.10 (0.09-0.11) | 0.97 (0.97-0.97) | 0.44 (0.41-0.49) | 0.74 (0.71-0.75) | 0.19 (0.18-0.21) | 0.91 (0.88-0.91) |
| | -1 to 0 years | | 0.56 (0.51-0.62) | 0.75 (0.74-0.75) | 0.10 (0.09-0.11) | 0.97 (0.97-0.97) | 0.70 (0.67-0.73) | 0.65 (0.64-0.66) | 0.25 (0.23-0.28) | 0.93 (0.92-0.94) |
| | -3 to -1 years | 0.5 | 0.11 (0.07-0.15) | 0.98 (0.97-0.98) | 0.19 (0.14-0.27) | 0.96 (0.95-0.97) | 0.07 (0.06-0.12) | 0.97 (0.96-0.97) | 0.23 (0.20-0.41) | 0.88 (0.87-0.90) |
| | -1 to 0 years | | 0.18 (0.16-0.24) | 0.96 (0.96-0.97) | 0.20 (0.18-0.27) | 0.96 (0.96-0.96) | 0.20 (0.15-0.25) | 0.94 (0.93-0.95) | 0.35 (0.31-0.44) | 0.88 (0.87-0.89) |
| AI-Echo | -3 to -1 years | 0.015 | 0.82 (0.79-0.87) | 0.53 (0.50-0.56) | 0.16 (0.14-0.20) | 0.96 (0.96-0.98) | 0.63 (0.52-0.73) | 0.64 (0.61-0.68) | 0.20 (0.19-0.23) | 0.92 (0.90-0.95) |
| | -1 to 0 years | | 0.91 (0.87-0.95) | 0.42 (0.40-0.44) | 0.15 (0.12-0.17) | 0.98 (0.96-0.99) | 0.86 (0.83-0.91) | 0.53 (0.51-0.54) | 0.27 (0.25-0.29) | 0.95 (0.94-0.97) |
| | -3 to -1 years | 0.05 | 0.78 (0.73-0.86) | 0.69 (0.67-0.73) | 0.22 (0.18-0.25) | 0.97 (0.96-0.98) | 0.49 (0.40-0.58) | 0.79 (0.76-0.82) | 0.25 (0.20-0.28) | 0.92 (0.90-0.94) |
| | -1 to 0 years | | 0.87 (0.82-0.92) | 0.62 (0.60-0.65) | 0.21 (0.17-0.23) | 0.98 (0.96-0.99) | 0.74 (0.68-0.78) | 0.68 (0.66-0.70) | 0.32 (0.30-0.36) | 0.93 (0.91-0.94) |
| | -3 to -1 years | 0.1 | 0.64 (0.59-0.71) | 0.82 (0.80-0.85) | 0.28 (0.22-0.35) | 0.95 (0.95-0.96) | 0.39 (0.36-0.45) | 0.86 (0.83-0.89) | 0.28 (0.23-0.35) | 0.91 (0.89-0.92) |
| | -1 to 0 years | | 0.80 (0.75-0.83) | 0.75 (0.74-0.77) | 0.27 (0.24-0.29) | 0.97 (0.96-0.98) | 0.70 (0.65-0.72) | 0.80 (0.78-0.82) | 0.42 (0.38-0.44) | 0.93 (0.91-0.94) |
| | -3 to -1 years | 0.25 | 0.36 (0.30-0.47) | 0.94 (0.93-0.96) | 0.40 (0.32-0.52) | 0.93 (0.92-0.95) | 0.25 (0.21-0.30) | 0.95 (0.93-0.97) | 0.43 (0.35-0.51) | 0.90 (0.88-0.91) |
| | -1 to 0 years | | 0.68 (0.65-0.74) | 0.91 (0.90-0.92) | 0.46 (0.42-0.51) | 0.96 (0.95-0.97) | 0.50 (0.44-0.54) | 0.93 (0.91-0.94) | 0.58 (0.51-0.64) | 0.90 (0.88-0.92) |
| | -3 to -1 years | 0.5 | 0.18 (0.11-0.23) | 0.99 (0.98-0.99) | 0.58 (0.44-0.74) | 0.92 (0.90-0.92) | 0.07 (0.03-0.15) | 0.99 (0.98-0.99) | 0.50 (0.21-0.69) | 0.88 (0.84-0.89) |
| | -1 to 0 years | | 0.35 (0.27-0.47) | 0.98 (0.97-0.99) | 0.71 (0.63-0.79) | 0.93 (0.92-0.94) | 0.26 (0.24-0.31) | 0.97 (0.96-0.98) | 0.65 (0.50-0.71) | 0.87 (0.86-0.88) |

AI: artificial intelligence; CI: confidence interval; ECG: electrocardiography; NPV: negative predictive value; PPV: positive predictive value; YNHHS: Yale-New Haven Health System.
The 95% confidence intervals are derived from bootstrapping with 1,000 replications.

**Table S5 | Sensitivity & specificity across timepoints using joint AI-Echo and AI-ECG testing.**

| | | | Yale-New Haven Health System (YNHHS)* | | | | | Houston Methodist Hospitals (HMH)** | | | | |
|---|---|---|---|---|---|---|---|---|---|---|---|---|
| **Method** | **Time period** | **Threshold** | **Positive screens (%)** | **Sensitivity (95% CI)** | **Specificity (95% CI)** | **PPV (95% CI)** | **NPV (95% CI)** | **Positive screens (%)** | **Sensitivity (95% CI)** | **Specificity (95% CI)** | **PPV (95% CI)** | **NPV (95% CI)** |
| AI-Echo **and** AI-ECG positive | -3 to -1 years | 0.015 | 151 (34.9) | 0.73 (0.57-0.88) | 0.68 (0.64-0.73) | 0.16 (0.10-0.22) | 0.97 (0.95-0.99) | 49 (28.2) | 0.67 (0.46-0.86) | 0.77 (0.70-0.84) | 0.29 (0.16-0.43) | 0.94 (0.90-0.98) |
| | -1 to 0 years | | 294 (44.8) | 0.78 (0.66-0.89) | 0.58 (0.54-0.62) | 0.13 (0.10-0.17) | 0.97 (0.95-0.99) | 242 (47.5) | 0.81 (0.73-0.89) | 0.60 (0.56-0.65) | 0.32 (0.27-0.38) | 0.93 (0.90-0.96) |
| | -3 to -1 years | 0.05 | 78 (18.0) | 0.61 (0.44-0.76) | 0.85 (0.82-0.89) | 0.26 (0.16-0.35) | 0.96 (0.94-0.98) | 26 (14.9) | 0.43 (0.22-0.65) | 0.89 (0.84-0.94) | 0.35 (0.17-0.53) | 0.92 (0.87-0.95) |
| | -1 to 0 years | | 174 (26.5) | 0.68 (0.53-0.80) | 0.77 (0.73-0.80) | 0.20 (0.14-0.25) | 0.97 (0.95-0.98) | 157 (30.8) | 0.67 (0.57-0.76) | 0.77 (0.73-0.81) | 0.41 (0.33-0.48) | 0.91 (0.88-0.94) |
| | -3 to -1 years | 0.1 | 41 (9.5) | 0.45 (0.27-0.62) | 0.94 (0.91-0.96) | 0.37 (0.23-0.52) | 0.95 (0.93-0.97) | 12 (6.9) | 0.19 (0.04-0.37) | 0.95 (0.91-0.98) | 0.33 (0.08-0.62) | 0.90 (0.85-0.94) |
| | -1 to 0 years | | 101 (15.4) | 0.56 (0.43-0.69) | 0.88 (0.85-0.91) | 0.28 (0.19-0.37) | 0.96 (0.94-0.97) | 104 (20.4) | 0.60 (0.50-0.70) | 0.89 (0.86-0.92) | 0.56 (0.46-0.65) | 0.91 (0.88-0.94) |
| | -3 to -1 years | 0.25 | 8 (1.8) | 0.09 (0.00-0.21) | 0.99 (0.98-1.00) | 0.38 (0.00-0.75) | 0.93 (0.90-0.95) | 7 (4.0) | 0.19 (0.05-0.38) | 0.98 (0.96-1.00) | 0.57 (0.17-1.00) | 0.90 (0.85-0.94) |
| | -1 to 0 years | | 43 (6.6) | 0.38 (0.25-0.51) | 0.96 (0.95-0.97) | 0.44 (0.28-0.59) | 0.95 (0.93-0.97) | 56 (11.0) | 0.43 (0.32-0.53) | 0.96 (0.94-0.98) | 0.73 (0.61-0.85) | 0.88 (0.85-0.91) |
| AI-Echo **or** AI-ECG positive | -3 to -1 years | 0.015 | 352 (81.3) | 0.97 (0.90-1.00) | 0.20 (0.16-0.24) | 0.09 (0.06-0.12) | 0.99 (0.96-1.00) | 141 (81.0) | 0.95 (0.85-1.00) | 0.21 (0.15-0.28) | 0.14 (0.09-0.20) | 0.97 (0.90-1.00) |
| | -1 to 0 years | | 552 (84.1) | 1.00 (1.00-1.00) | 0.17 (0.14-0.20) | 0.09 (0.07-0.12) | 1.00 (1.00-1.00) | 444 (87.2) | 0.98 (0.95-1.00) | 0.15 (0.12-0.19) | 0.21 (0.17-0.25) | 0.97 (0.92-1.00) |
| | -3 to -1 years | 0.05 | 269 (62.1) | 0.91 (0.81-1.00) | 0.40 (0.35-0.45) | 0.11 (0.07-0.15) | 0.98 (0.96-1.00) | 108 (62.1) | 0.86 (0.69-1.00) | 0.41 (0.34-0.50) | 0.17 (0.10-0.24) | 0.95 (0.90-1.00) |
| | -1 to 0 years | | 449 (68.4) | 0.94 (0.86-1.00) | 0.34 (0.30-0.38) | 0.10 (0.08-0.14) | 0.99 (0.97-1.00) | 383 (75.2) | 0.94 (0.89-0.98) | 0.29 (0.25-0.33) | 0.23 (0.19-0.28) | 0.95 (0.91-0.99) |
| | -3 to -1 years | 0.1 | 203 (46.9) | 0.91 (0.80-1.00) | 0.57 (0.52-0.62) | 0.15 (0.10-0.20) | 0.99 (0.97-1.00) | 87 (50.0) | 0.81 (0.62-0.95) | 0.54 (0.46-0.62) | 0.20 (0.11-0.28) | 0.95 (0.91-0.99) |
| | -1 to 0 years | | 362 (55.2) | 0.88 (0.78-0.96) | 0.48 (0.44-0.52) | 0.12 (0.09-0.16) | 0.98 (0.96-0.99) | 328 (64.4) | 0.89 (0.82-0.94) | 0.41 (0.37-0.46) | 0.26 (0.21-0.31) | 0.94 (0.90-0.97) |
| | -3 to -1 years | 0.25 | 100 (23.1) | 0.58 (0.42-0.75) | 0.80 (0.76-0.84) | 0.19 (0.12-0.26) | 0.96 (0.93-0.98) | 50 (28.7) | 0.48 (0.27-0.71) | 0.74 (0.66-0.81) | 0.20 (0.10-0.32) | 0.91 (0.86-0.96) |
| | -1 to 0 years | | 220 (33.5) | 0.80 (0.69-0.91) | 0.70 (0.67-0.74) | 0.18 (0.13-0.24) | 0.98 (0.96-0.99) | 234 (46.0) | 0.77 (0.68-0.85) | 0.61 (0.57-0.66) | 0.32 (0.25-0.38) | 0.92 (0.89-0.95) |

*In YNHHS, the denominator includes 433 and 656 unique participants with both AI-Echo and AI-ECG predictions in the "-3 to -1 years" and "-1 to 0 year" period, respectively.
** In HMH, the denominator includes 174 and 509 unique participants with both AI-Echo and AI-ECG predictions in the "-3 to -1 years" and "-1 to 0 year" period, respectively
AI: artificial intelligence; CI: confidence interval; ECG: electrocardiography; NPV: negative predictive value; PPV: positive predictive value; YNHHS: Yale-New Haven Health System.
The 95% confidence intervals are derived from bootstrapping with 1,000 replications.

### Supplemental Figures

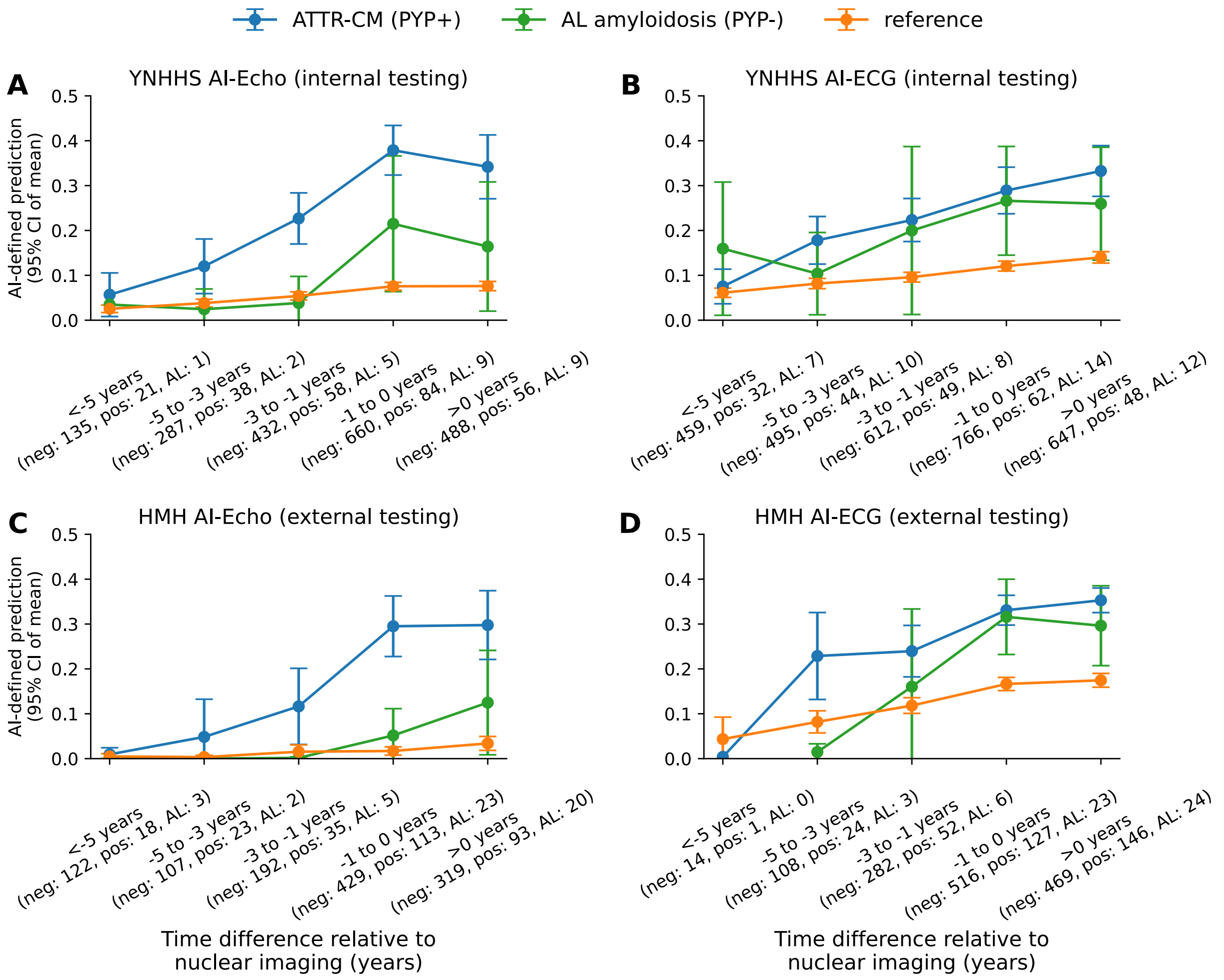


**Supplemental Figure S1 | Evolution of AI-Echo and AI-ECG ATTR-CM probabilities across subgroups of light chain (AL)-positive and nuclear cardiac amyloid imaging-positive cases.** The panels illustrate the mean (with error bars denoting the 95% confidence interval of mean) of the AI-Echo and AI-ECG-derived probabilities for ATTR-CM for individuals split across three strata: (i) those who tested positive by nuclear cardiac amyloid imaging (blue); (ii) those who tested negative but received a diagnosis of AL amyloidosis (green); and (iii) a reference sample of controls with negative nuclear imaging and no AL amyloidosis diagnosis code. The x axis denotes the time between the TTE/ECG and the timing of nuclear cardiac amyloid testing summarized across discrete time groups (negative time differences suggest that the TTE/ECG was performed before the nuclear cardiology exam). The brackets below each period along the x axis denote the number of unique positive and negative individuals in each period for each one of the three classes. Results are presented across YNHHS (internal cohort; A-B) and HMH (external cohort; C-D). If more than one study was available in a discrete time period for an individual participant, we used the median of all predictions for that participant in that time window. ATTR-CM: transthyretin amyloid cardiomyopathy; ECG: electrocardiography; HMH: Houston Methodist Hospitals; TTE: transthoracic echocardiography; YNHHS: Yale-New Haven Health System.

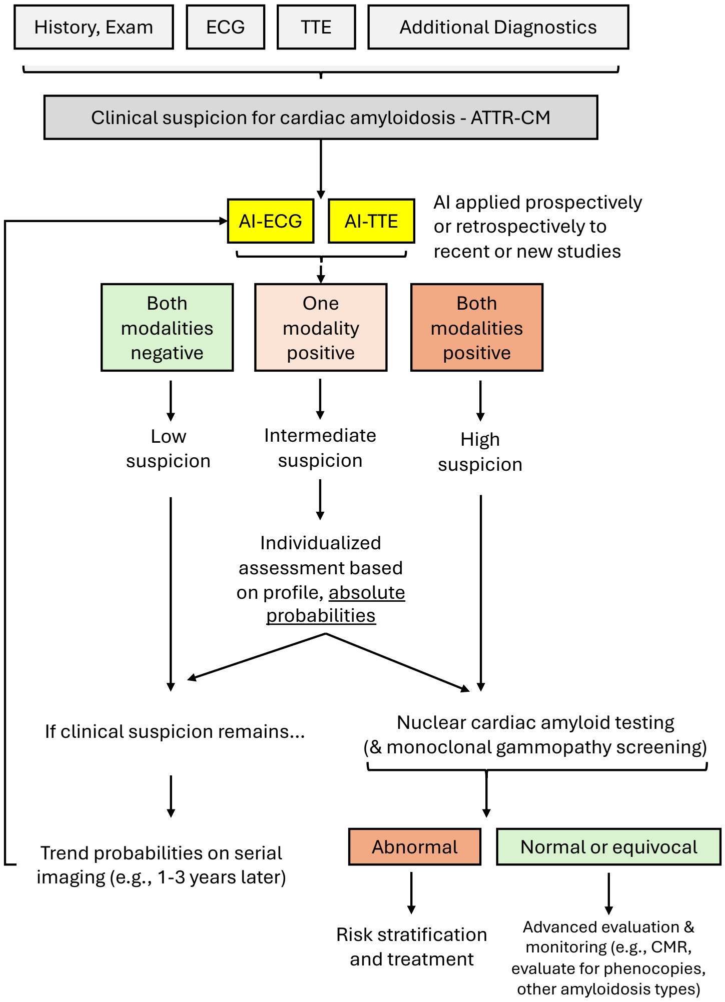


**Supplemental Figure S2 | Example of a possible algorithm for the integration of AI-Echo and AI-ECG in the screening and diagnosis of ATTR-CM.** AI can be applied to both existing and prospectively acquired TTEs and ECGs to screen for the presence of ATTR-CM. If several studies have been performed in the past, analysis can be limited to the more recent exam, although longitudinal analysis of time trends can provide insights into possible disease onset and progression. Generally, at sensitive thresholds (e.g., 1.5%, or 5%) double-negative screening on both modalities confers high sensitivity, whereas double-positive screening confers high specificity and may enable efficient triage of downstream nuclear cardiac amyloid imaging and further testing. Discrepant results (one modality, one negative) may warrant further evaluation based on clinical history, degree of suspicion, and examining the absolute probability of each modality that represents a possible surrogate of model confidence. This pathway is hypothesis-generating and should not be used to inform clinical practice without prospective validation in prospective clinical trials. AI: artificial intelligence; ATTR-CM: transthyretin amyloid cardiomyopathy; CMR: cardiac magnetic resonance imaging; ECG: electrocardiography; TTE: transthoracic echocardiography.
